# Equivalent Gain-of-Function Variants in *KCNK3* and *KCNK9* and their Contribution to Distinct TASK K2P Channelopathies

**DOI:** 10.64898/2025.12.15.25342159

**Authors:** Kate M. Crowther, Thibault R.H. Jouen-Tachoire, Peter Proks, Peter Rory Hall, Emma L. Veale, Janina Sörmann, Karin E.J. Rödström, Thomas Müller, Saskia B. Wortmann, Nina Barisic, Natalie Hauser, Vincenzo Salpietro Damiano, RaeLynn Forsyth, Linford Williams, Carlos A. Bacino, Jill A Rosenfeld, Henry Houlden, Simon Newstead, Caroline F. Wright, James Fasham, Alistair A. Mathie, Reza Maroofian, Stephen J. Tucker

## Abstract

Gain-of-function (GoF) missense variants in the Two-Pore Domain (K2P) K^+^ channel TASK-1 (*KCNK3*) result in Developmental Delay with Sleep Apnea (DDSA), a neurodevelopmental channelopathy, whilst loss-of-function (LoF) variants cause a hypertensive disorder. However, for the related TASK-3 channel (*KCNK9*), both LoF and GoF variants underlie a distinct neurodevelopmental disorder, *KCNK9* Imprinting Syndrome (KIS). The relationship between genotype and phenotype in these disorders is further complicated because TASK-1 and TASK-3 can co-assemble into heteromeric channels with distinct functional properties. Here we report additional probands with missense variants in *KCNK3* and *KCNK9* and investigate the effect of four novel variants on the functional properties of both homomeric and heteromeric TASK channels. Interestingly, two of these new GoF variants (R131H and L122V) are found in both TASK-1 and TASK-3 and have equivalent functional effects on heteromeric TASK-1/TASK-3 channels, yet result in different clinical phenotypes. We have also determined a cryoEM structure for the pathogenic L122V mutant TASK-3 channel which suggests its dramatic functional effect is likely due to subtle changes in gating and permeation within the inner cavity. Overall, these results highlight the dominant role that homomeric TASK channels play in defining their associated channelopathies as well as the complexity of interpreting K^+^ channel dysfunction in pathophysiology.

## Introduction

Variants that affect the function of K2P K^+^ channels are now increasingly associated with a range of diseases including diabetes, nephropathy, migraine, sleep apnea, hypertension, and other cardiac disorders, as well as a range of neurodevelopmental disorders^1^. However, despite the pressing need, there are few effective treatments for many of these complex conditions, and their underlying mechanisms are often not well understood.

There are 15 different human K2P (*KCNK*) channels and this family are often referred to as simple ‘leak’ conductances involved in establishing the resting membrane potential, but it is now clear they exhibit complex patterns of dynamic regulation by diverse stimuli, including many GPCR-associated regulatory pathways^2, 3^. Understanding K2P channel dysfunction in the disease state is therefore critical to better understanding these rare channelopathies and to exploit their therapeutic potential in other more common disease states^4, 5^.

K2P channels represent a structurally distinct subfamily of K^+^ channels. In addition to their distinct extracellular Cap domain, each subunit contains four transmembrane helices (M1-M4) and two pore domains (P1 and P2). Two subunits therefore assemble as a ‘dimer of dimers’ to create a pseudotetrameric channel that is similar to other classical K^+^ channel pores^3^.

*KCNK3* and *KCNK9* encode the TASK-1 and TASK-3 channels respectively. Inherited LoF variants in *KCNK3* are associated with a form of pulmonary arterial hypertension (PAH)^6, 7, 8^ and we have recently shown that *de novo* heterozygous missense GoF variants underlie a novel neurodevelopmental disorder (DDSA) where all of those affected also have sleep apnea^9, 10^.

TASK-1 is expressed in many cells and tissues involved in the control of breathing/ventilation including the carotid body, lung, heart, and pulmonary artery smooth muscle, as well as many ventilatory-associated chemosensitive regions of the brain, hypoglossal motor neurons, and spinal cord motor neurons^11, 12, 13^. They are also a target for volatile anaesthetics^14, 15^, and inhibitors that target TASK-1 are in development for the treatment of sleep apnea with improvements observed in a subset of patients where topical application of inhibitor in the upper airway was achieved via nasal spray^16^.

The *KCNK3* GoF variants associated with DDSA all produce TASK-1 channels with an intrinsically higher basal activity and markedly reduced sensitivity to inhibition by Gq-protein coupled receptor (GqPCR) pathways^9^. Structural analysis reveals that the majority of these mutations cluster in or around regions important for channel gating in the M2 and M4 transmembrane helices. In particular, these mutations are found near an intersection of these helices which forms the ‘X-gate’, a structural motif which is analogous to the classical bundle-crossing gate in other K^+^ channels^9, 17^. Indeed, we have shown that one of the recurrent *KCNK3* variants, N133S, disrupts interactions between the M2 and M4 helices that stabilise the X-gate in its closed conformation resulting in channels with a >20-fold increase in single-channel open probability (*P_o_*).

Missense variants in the related TASK-3 channel (*KCNK9*) are also associated with a different neurodevelopmental disorder, ‘*KCNK9* Imprinting Syndrome’ (KIS), previously known as Birk-Barel syndrome^18, 19^. Typically, KIS-associated mutations produce a LoF in TASK-3, but more recently, GoF mutations have also been reported in probands that present similar phenotypes^20^. TASK-3 exhibits a high degree of homology with TASK-1 sharing 54% overall amino acid identity and almost 80% identity within the pore-forming transmembrane regions. Consequently, as well as exhibiting significant structural similarity, their functional properties share many common features^17, 21, 22^. But although sleep apnea has been reported in some KIS probands^20^, it is not a universal feature, unlike its defining phenotype with *KCNK3* GoF variants in DDSA.

It remains unclear why both LoF and GoF variants in *KCNK9* result in the same KIS phenotype, but this complex aetiology may be due to the paternal imprinting of *KCNK9* and mosaicism, as well as the major changes that result from the inability to regulate mutant channel activity that is common to GoF mutations. Another complicating factor in tissues where *KCNK3* and *KCNK9* expression overlaps, is the ability of TASK channels to coassemble to form novel heteromeric channels with unique functional properties^23, 24, 25, 26^. For example, heteromeric TASK-1/TASK-3 channels form the major oxygen-sensitive background K^+^ channel in carotid body glomus cells which plays a critical role in the regulation of respiration^27, 28^. Equivalent mutations in either channel subunit might therefore be expected to influence these currents in similar ways, but their relative effect and their contribution to these different disorders remains unclear.

In this study we identify eight new probands with GoF mutations in TASK-1 which display a DDSA phenotype, including three novel *de novo* missense variants (F125S, Q126E, and R131H). We also functionally and structurally characterise a novel missense variant in TASK-3 (L122V) associated with KIS and show how these mutations provide important insights into the relative contributions of homomeric and heteromeric TASK channels in the development of TASK-related disorders.

## Methods

### Patient identification, genetic and clinical investigation, and ethical approval

Using the GeneMatcher platform^10^ and extensive international data sharing, we identified nine families. A uniform clinical pro forma was distributed to collect clinical details. Parents or legal guardians of all affected individuals provided informed consent for the publication of clinical information and genetic data, in accordance with the Declaration of Helsinki. The study was approved by the relevant local Ethics Committees, including University College London (for Integrated Research Application System (IRAS) Project ID: 310045) whilst the original projects^9, 10^ have UK Research Ethics Committee approval (10/H0305/83, granted by the Cambridge South REC, and GEN/284/12 granted by the Republic of Ireland REC). Trio exome or genome sequencing, as well as proband-only sequencing where applicable, was performed on DNA extracted from blood-derived leukocytes. Data analysis and variant filtration were conducted using standard pipelines, and Sanger sequencing was used for segregation analysis when indicated.

### Electrophysiology

This was done as previously described^9^. Briefly, the wild-type human TASK-1 gene (*KCNK3*) and human TASK-3 gene (*KCNK9*) were subcloned into pFAW. Mutations were introduced by site-directed mutagenesis and confirmed by sequencing. mRNA was transcribed using the T7 mScript^TM^ Standard mRNA Production System. Each oocyte was then injected with 2 ng of RNA and incubated for 20–24 h at 17.5°C. Recordings were performed in ND96 buffer at pH 7.4 (96 mM NaCl, 2 mM KCl, 2 mM MgCl_2_, 1.8 mM CaCl_2_, 5 mM HEPES). Currents were recorded using a 400 ms voltage step protocol from a holding potential of -80 mV delivered in 10 mV increments between -120 mV and +50 mV and 800 ms ramp protocols from -120 to +50 mV. All recorded traces were analysed using Clampfit, and graphs were plotted using Origin2021. Single-channel currents in cell-attached patches were recorded in symmetrical 140 mM KCl solutions containing 10 mM HEPES (pH 7.4 with KOH). Data was filtered at 2 or 5 kHz and recorded at a 200 kHz sampling rate with program Clampex on an Axopatch 200B amplifier. Data analysis was performed using Clampfit. Channel *P_o_* was determined from single-channel recordings with 1-4 min duration which exhibited only one main open level.

### Protein Expression and purification

Overlapping mutagenesis PCR was conducted to generate the TASK-3 L122V mutant. TASK-3 L122V was purified as stated by Hall et al., 2024^20^.

### Cryo-EM sample preparation and collection

Samples were prepared as previously described^21^; briefly, TASK-3 L122V (4.0 mg/mL) was applied onto glow-discharged AuFlat 300 1.2/1.3 grids (Sigma-Aldrich). The grids were blotted for 4–5 seconds at 100% humidity and 4°C, then vitrified in liquid ethane using a Vitrobot Mark IV (Thermo Fisher Scientific). Data was collected on a Titan Krios G3 (FEI) operating at 300 kV, equipped with a BioQuantum imaging filter (Gatan) and a K3 direct detection camera (Gatan). Images were acquired at a magnification of 105,000×, corresponding to a physical pixel size of 0.832 Å with a total dose rate of 43.9 e⁻/Å². A total of 13,035 initial movies were collected.

### Image processing

All processing was performed in cryoSPARC^29^ unless stated otherwise. Movies were imported and underwent motion correction and patched contrast transfer function (CTF) estimation. A subset of 1,531 micrographs was selected for blob picking with a particle diameter of 125 Å. The initially extracted particles underwent two rounds of 2D classification and *ab initio* reconstruction (n=2), which yielded a volume used to generate templates for particle picking.

The TASK-3 L122V templates were used for particle repicking from the full dataset with a diameter of 125 Å, resulting in the extraction of 8,381,007 particles with box size of 256 px, Fourier cropped to 128 px. These particles underwent four rounds of 2D classification during which junk particles were removed, leaving 1,718,239 particles. The particles were re-extracted with a full 256 px box size, prior to two more rounds of 2D classification. Two rounds of five *ab initio* classes and heterogenous refinement in C1 and C2 symmetry were completed leading to a particle stack of 332,112 particles. Homogenous refinement, non-uniform (NU) refinement and reference-based motion correction in C2 produced a 2.90 Å map. Polished particles were processed with cryosieve^30^, and the particle stacks were re-imported into cryoSPARC. To identify the highest resolution stack, each stack was subjected to one round of 2D classification, *ab initio* reconstruction in C1, followed by homogeneous and NU refinements in C2. The final particle set (211,335 particles) produced a map with a resolution of 2.83 Å, based on the FSC = 0.143 criterion.

### Model building and refinement

The initial model for TASK-3 L122V (PDB ID: 9G9V) was obtained from the Protein Data Bank and manually mutated and adjusted within the map using Coot. The model was further refined iteratively, first through manual adjustments in Coot and then using phenix.real_space_refine^31^. The refined model was subsequently processed with ISOLDE^32^ (through ChimeraX^33^) to enhance their accuracy and underwent a final round of refinement in Phenix, with the ISOLDE-optimized models serving as references. Structural figures were generated using PyMOL (Schrödinger, LLC).

## Results

### Eight new DDSA probands including three with novel variants in KCNK3

Our previous study identified nine probands with DDSA^9^, each heterozygous for one of six *de novo* missense variants in *KCNK3*. Since that original report in 2022 we have now identified an additional five DDSA probands with similar *de novo* GoF mutations in *KCNK3,* including four with the recurrent N133S variant, and a further proband with the L241F variant. In addition, we have identified three further probands with novel heterozygous missense variants in *KCNK3* who all match the reported DDSA neurodevelopmental phenotype (**Supplementary Table 1**). Interestingly, the three new variants (F125S, Q126E, and R131H) all cluster in the M2 transmembrane helix adjacent to several previously characterised DDSA variants (**Figure 1a**). The location of these novel variants alongside others within M2 indicates that this particular region represents a major hotspot for pathogenic mutations and is consistent with the reported effects of other mutations within this region on K2P channel activity.

**Figure 1:**
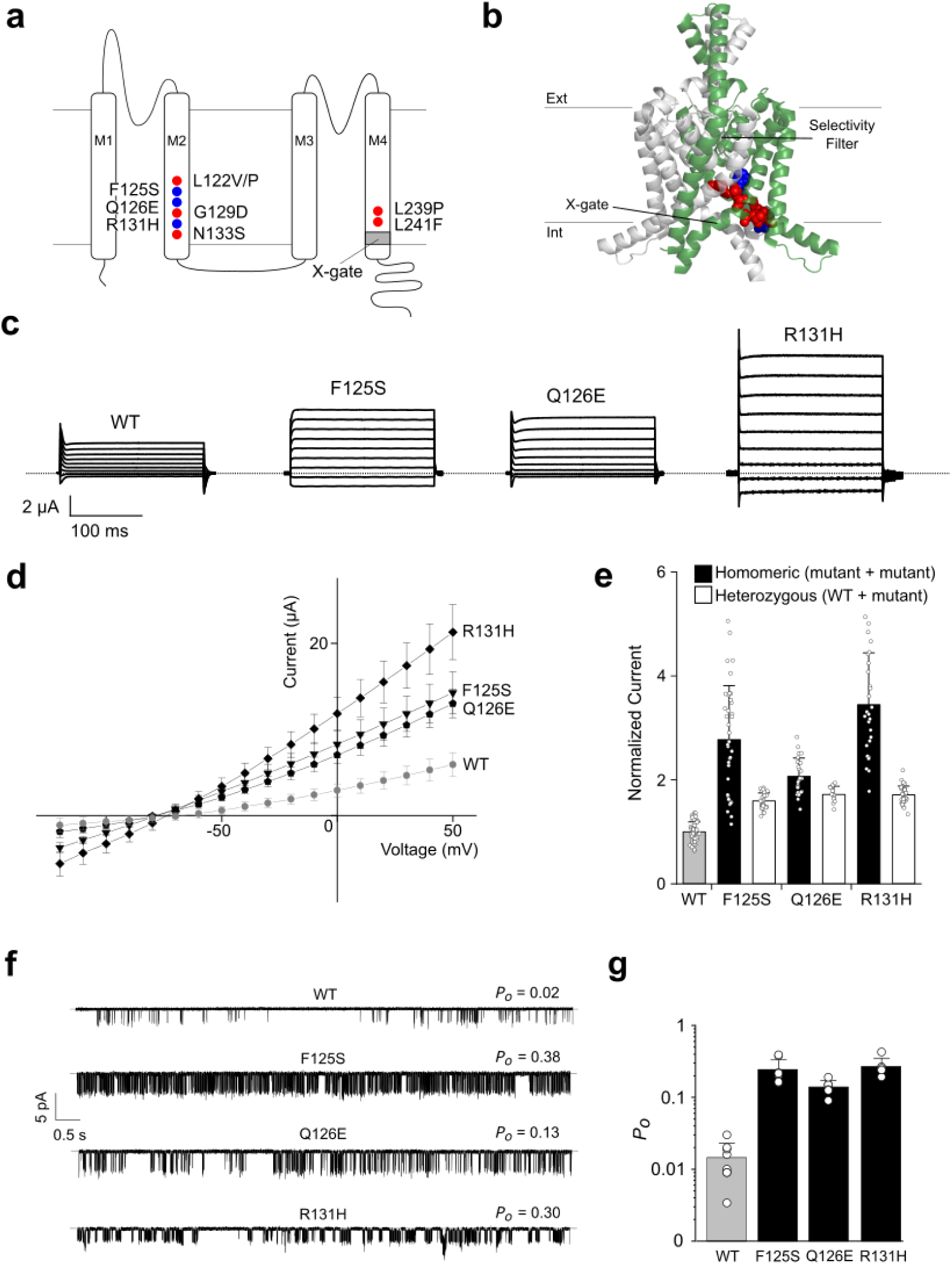
Functional effects of new DDSA-associated variants. (**a**) 2D topological model of a TASK-1 subunit with the position of the new and previously characterised DDSA variants labelled in blue and red respectively. The X-gate is labelled in dark grey. (**b**) TASK-1 crystal structure (PDB: 6RV2). Subunits are shown in green and grey respectively. The new and previously-characterised DDSA mutations are shown as blue and red spheres respectively. (**c**) Representative TEVC recordings of WT TASK-1 and DDSA mutant currents in response to voltage steps from −120 to +50 mV in 20 mV steps from a holding potential of −80 mV. (**d**) Current–voltage plot of WT TASK-1 (*n* = 8), F125S (*n* = 16), Q126E (*n* = 19), R131H (*n* = 23); data are presented as mean ± s.d. (**e**) Currents for homomeric DDSA mutants, and ‘heterozygous’ channels formed from 1:1 coexpression of WT TASK-1 and DDSA mutants normalized to WT current at +50 mV: WT (*n* = 43), F125S (*n* = 26), F125S-WT (*n* = 16), Q126E (*n* = 22), Q126E-WT (*n* = 12), R131H (*n* = 17), R131H-WT (*n* = 23) data are presented as mean ± s.d. All mutant currents differ from WT (*P* < 0.01, two-tailed Student’s *t*-test). (**f**) Representative single-channel recordings of new DDSA mutants at a holding potential of −160 mV. (**g**) Single-channel *P_o_* values at –100 mV for each mutant. WT (*n* =8), F125S (*n* = 8), Q126E (*n* =6), R131H (*n* =6). All mutant *P_o_* values differ from that for WT (*P* < 0.01).

### Novel DDSA variants also produce GoF channels with impaired GPCR-sensitivity

All existing DDSA variants are associated with a GqPCR-insensitive GoF effect on TASK-1 channel activity. Therefore, to confirm these new DDSA variants also fit this profile we next examined their functional activity. Whole-cell K^+^ currents of wild-type (WT) and mutant TASK-1 channels were measured by two-electrode voltage clamp (TEVC) after expression in *Xenopus laevis* oocytes. These results show that all three variants produced markedly larger currents than WT TASK-1 (**Figure 1**).

K2P channels assemble as dimers and each of the new DDSA probands are also heterozygous for these *KCNK3* variants. To better replicate this we therefore also measured average whole-cell currents from oocytes co-injected with equal amounts of WT and mutant TASK-1 RNA. Similar to the results obtained with the previous DDSA variants, we found that all three novel variants also increased these ‘heterozygous’ currents by approximately 50% (**Figure 1e**).

To confirm that these increases are due to an increase in the intrinsic activity of the channel rather than a major increase in trafficking to the cell surface we also measured single channel currents of homomeric WT and mutant TASK-1 channels and found that all three variants resulted in a marked (∼10-fold) increase in channel open probability (**Figure 1f,g**). A modest (20%) increase in single channel conductance was also observed for the F125S and Q126E variants, but this was not large enough to account for their increased whole-cell currents (>250%).

WT TASK-1 channels are sensitive to inhibition by GqPCR mediated pathways and existing DDSA variants are characterised by a dramatic reduction in this sensitivity that exacerbates their intrinsic GoF effect by uncoupling them from their regulatory pathways^9^. We therefore also examined the GqPCR-sensitivity of these new variants and found that whereas ATP-induced stimulation of a coexpressed P2Y2 Receptor results in ∼50% inhibition of WT TASK-1 currents, all three variants exhibited a marked reduction with only 15-20% inhibition observed in identical conditions. This effect was seen in both homomeric and’heterozygous’ TASK-1 mutant channels (**Figure 2)**.

**Figure 2:**
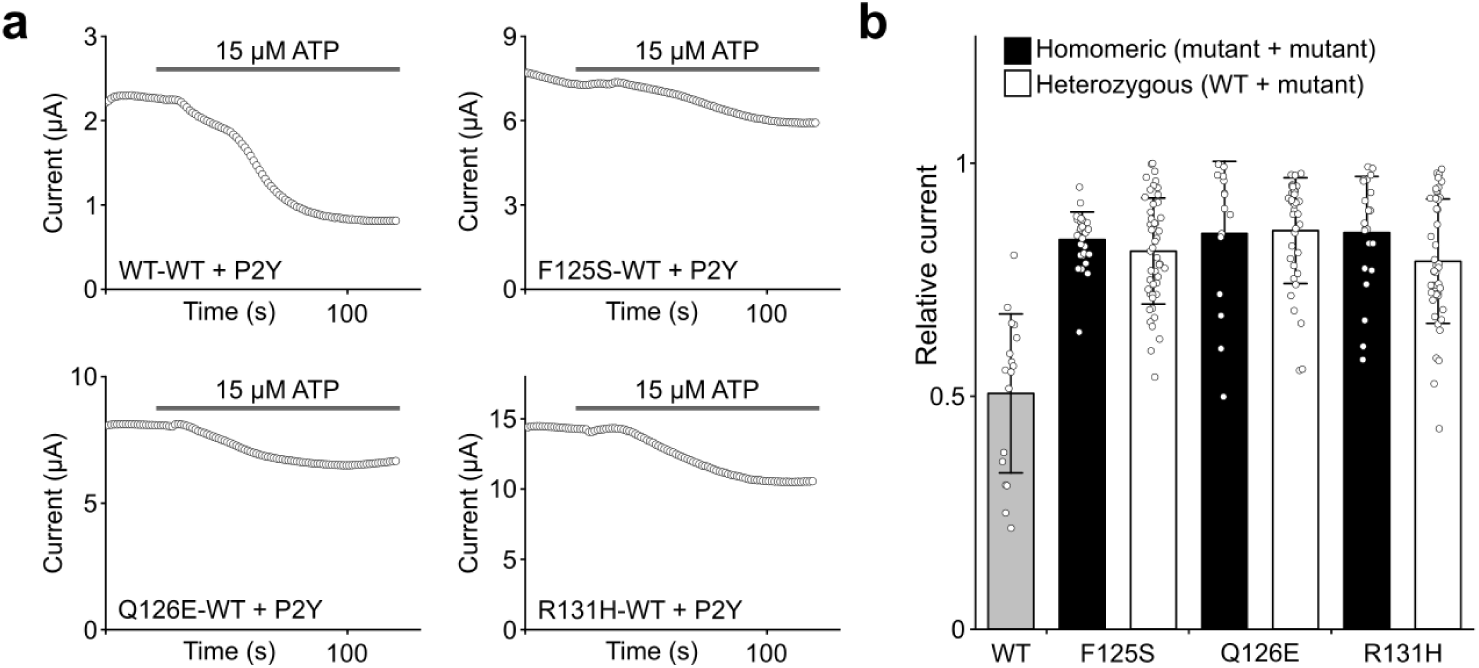
GqPCR-mediated inhibition is reduced by DDSA-associated mutations. (**a**) Representative currents at +50 mV of WT TASK-1 channels (WT-WT) and ‘heterozygous’ channels from coexpressed WT and mutant subunits, over time while adding 15 µM ATP. This concentration produces ∼50% inhibition of WT TASK-1. (**b**) Residual current for homozygous mutants and mutants coexpressed 1:1 with WT TASK-1 after addition of 15 μM ATP. WT TASK-1 (*n* = 17), F125S (*n* = 28), F125S/WT (*n* = 48), Q126E (*n* = 15), Q126E/WT (*n* = 36), R131H (*n* = 22), R131H/WT (*n* = 45); data are presented as mean ± s.d. All mutant currents differ from WT (*P* < 0.01, two-tailed Student’s *t*-test).

Together, these results confirm that these three novel *KCNK3* variants also fit same functional profile for other DDSA variants by producing channels with an increased activity that is markedly less sensitive to inhibition via GqPCR-coupled signalling pathways. These functional defects will therefore have profound effects on the electrical activity and excitability of cells in which *KCNK3* is expressed.

An additional inherited variant in *KCNK3* was also identified in a proband with behavioural problems later diagnosed as an autism spectrum like disorder, but with no sleep apnea. This variant (T121M) is located in the M2 helix adjacent to the pathogenic L122V DDSA variant. However, functional analysis revealed this variant to behave similar to WT TASK-1 producing equivalent levels of whole-cell currents with no increased single channel open-probability and only a mild reduction in GqPCR sensitivity (**Supplementary Figure S1**). Thus, although the M2 helix clearly represents a hotspot for DDSA mutations, this variant is therefore unlikely to underlie the phenotype observed in this proband. It also highlights the structural boundary of this specific hotspot for GoF variants.

### Equivalent disease-causing mutations in both TASK-1 and TASK-3

Due to their high degree of sequence homology, TASK-1 and TASK-3 share many structural and functional properties, and we have previously shown that GoF variants found in DDSA produce identical functional effects when equivalent mutations are introduced at the same sites in TASK-3 channels^9^.

Interestingly, the R131H DDSA variant reported above has also now been identified as a disease-causing GoF variant in TASK-3 where it is associated with KIS^20^. This therefore suggests that a pathogenic GoF variant in TASK-1 also has the potential to cause disease if the equivalent mutation occurs in TASK-3, and vice-versa. In support of this, the L122V variant not only results in DDSA in TASK-1 (*KCNK3*), but we also now report here that it is associated with KIS when mutated in TASK-3 (*KCNK9*) as we identified a proband who fits the classical KIS developmental phenotype with a maternally-inherited L122V variant in *KCNK9* (**Supplementary Table 1**).

Mutations at this particular site (TM2.6) in the M2 helix cause an activatory GoF effect in almost every known K2P channel^34^, and in TASK-1 produce channels with increased channel open-probability and reduced GqPCR-sensitivity^9^. We therefore examined its effect in TASK-3 and found similar activatory effects in both homomeric TASK-3 (L122V/L122V) and heterozygous (WT/L122V) channels. As in TASK-1, this GoF is due to an increased single channel open-probability (*P_o_*) (**Figure 3**).

**Figure 3:**
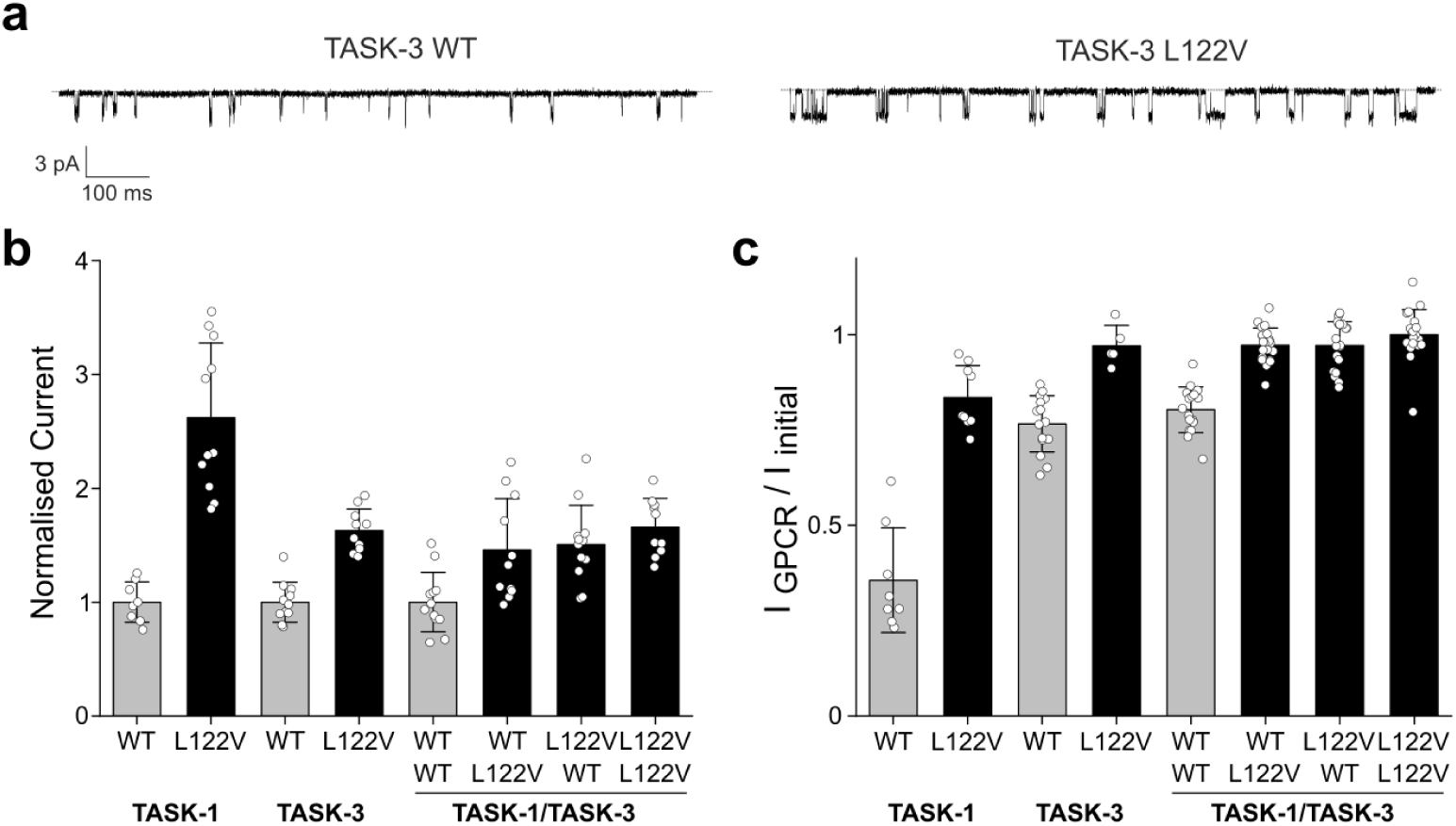
Position-independent effect of a single L122V mutation in heteromeric TASK channels. (**a**) Recordings of single channel currents at –160 mV showing the marked increase in *P_o_* for L122V TASK-3 channels compared to WT. (**b**) Currents for homomeric and heteromeric WT and L122V TASK channels normalized to WT current at +50 mV. For heteromeric TASK-1/TASK-3 channels: top row = TASK-1, bottom = TASK-3. *n* > 10 for all conditions. (**c**) Residual currents for homomeric and heteromeric TASK channels with different combinations of L122V mutations as indicated after addition of 15 μM ATP. Labels same as shown in panel b. *n* > 10 for all conditions; data are presented as mean ± s.d. All mutant currents differ from WT (*P* < 0.01, two-tailed Student’s *t*-test).

The dominant functional effect of this variant even in heterozygous TASK-3 channels not only fits with its pattern of inheritance, but also suggests that it may have an equivalent activatory effect on heteromeric TASK-1/TASK-3 channels, similar to the effect of the TASK-1 L122V variant.

We therefore examined the functional effects of the L122V variant on heteromeric TASK-1/TASK-3 channel activity and found that whole-cell currents were increased ∼50% along with a markedly impaired sensitivity to GqPCR-inhibition irrespective of which TASK subunit contained the mutation **(Figure 3b).**

### Structural consequences of the TASK-3 L122V variant

The common activatory effect of TM2.6 mutations on K2P channel activity suggests a conserved gating mechanism in this region^34^. In a previous study we demonstrated that hydrophilic substitutions at this position reduced a hydrophobic barrier deep within the inner cavity of TWIK-1 (*KCNK1*) ^35^, but it remains unclear whether this is the case in all other K2P channels.

We have recently determined a structure of the WT TASK-3 channel by single-particle cryoEM^21^ and so used an identical method to determine the structure of this L122V variant to 2.8 Å resolution. Comparison of this structure with WT TASK-3 reveals an identical overall fold *i.e.*, the channel is in the closed X-gate conformation with no obvious visible differences except within the inner cavity at the site of the mutation. In WT TASK-3 the closed X-gate forms a small inner cavity just below the filter with the L122 side-chain lining this cavity (**Figure 4 and Supplementary Figure S2**). In the L122V mutant structure the smaller nature of the side chain increases the radius of the pore at this point from 3 to 4 Å and thus also increases the overall volume of the cavity immediately below the filter. This change is therefore perfectly situated to affect the interaction between K^+^ and water immediately prior to entry of K^+^ ions into the filter from within the inner cavity, but it may also have allosteric effects on the filter gate itself.

**Figure 4:**
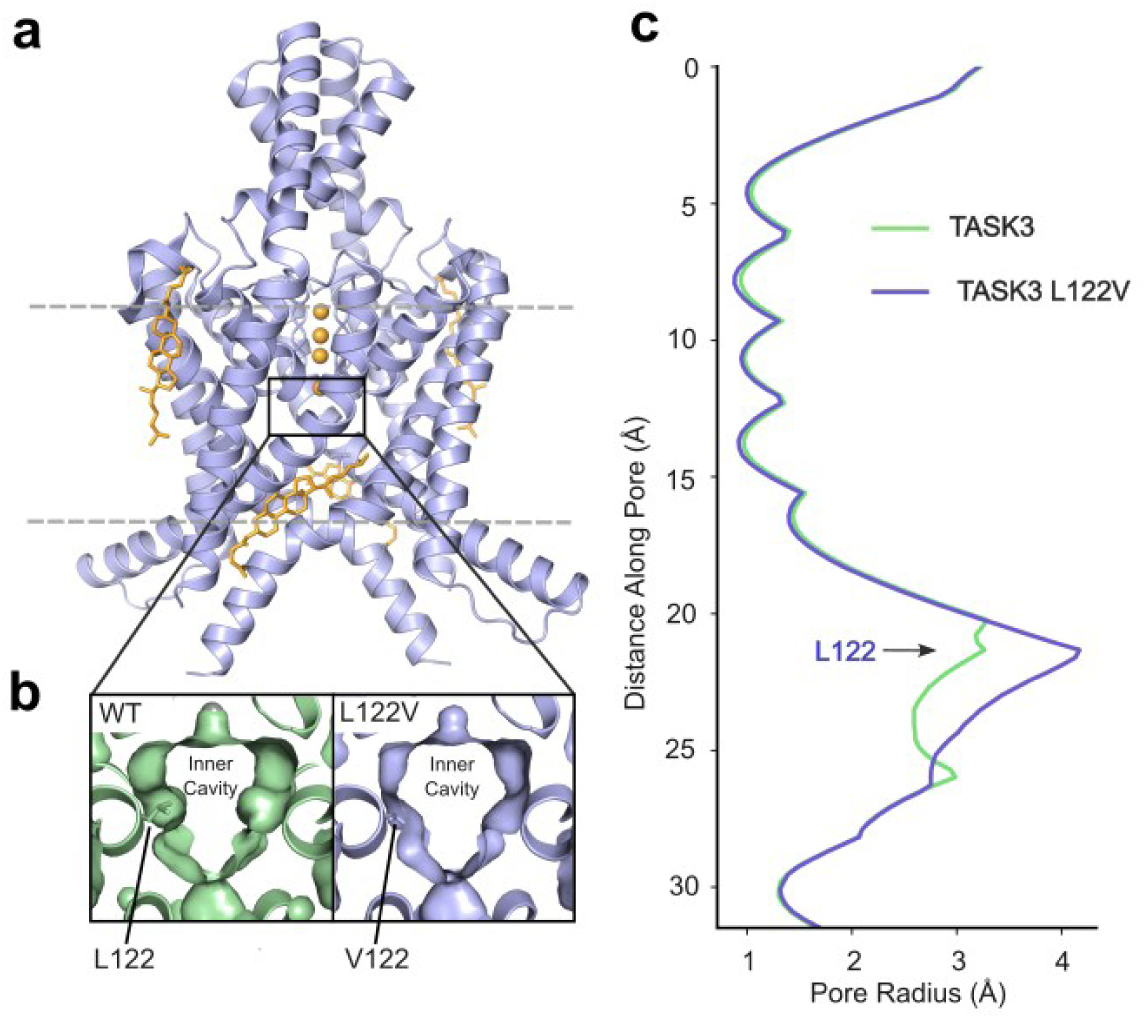
Cryo-EM structure of the TASK-3 L122V variant reveals expansion of inner cavity below filter. (**a**) Cartoon representation of the overall fold of the TASK-3 L122V mutant channel. K^+^ ions are shown as gold spheres and cholesterol hemisuccinate molecules as gold sticks. (**b**) Expanded view of the inner cavity at the site of the L122V mutation shown as a surface representation. The mutant L122V is shown in purple and WT TASK-3 in green. The side chains at position 122 are shown as sticks. Note the expanded inner cavity in the mutant channel (**c**) Pore radius plot of WT TASK-3 and TASK-3 L122V – the expansion of the inner cavity below the filter at position 122 is indicated.

## Discussion

In this study, we have identified five further DDSA probands with known variants in *KCNK3,* and functionally characterised three additional *de novo* heterozygous missense variants (F125S, Q126E, R131H) associated with DDSA. This markedly expands the global cohort of individuals with DDSA and reinforces the role that defective TASK-1 channels play in this disorder. We have also functionally and structurally characterised a novel variant in TASK-3 (L122V) associated with KIS. These variants all share similar GoF effects on channel activity with a markedly reduced sensitivity to inhibition via GqPCR-coupled pathways. These GoF effects and regulatory defects were also dominant in heterozygous channels containing one mutant subunit as well as in heteromeric TASK-1/TASK-3 channels. Furthermore, we show that two of these pathogenic mutations (R131H and L122V) are found in both *KCNK3* (TASK-1) and *KCNK9* (TASK-3) channels, and that their effects on heteromeric TASK channel activity appear the same, irrespective of which channel subunit contains the mutation.

Many subfamily members within the large superfamily of tetrameric cation channels share structural and functional similarities and examples already exist of equivalent mutations in domains critical for channel function that result in similar functional defects, yet different phenotypes. For example, the mutation of equivalent residues within the highly conserved DIV S3-S4 linker of the skeletal and cardiac muscle voltage-gated sodium channels (*SCN4A* and *SCN5A* respectively) produce similar effects on channel inactivation in these channels, yet result in different channelopathies due to the distinct spatiotemporal expression of these two channels^36^. The difference with the equivalent TASK channel mutations (L122V and R131H) examined here is the overlapping expression patterns of TASK-1 and TASK-3 channels and the ability of these two subunits to form physiologically relevant heteromeric channels in these cell types. In such cases any differences in the activity of heteromeric channels produced by these mutations become highly significant.

However, our results show that these equivalent mutations in either TASK-1 or TASK-3 have very similar effects, not just on homomeric but also heteromeric channel activity. Given the very high degree of structural and functional conservation between TASK-1 and TASK-3, this is perhaps not surprising. However, the markedly different disease phenotypes produced by these mutations therefore suggests it is not defective heteromeric channel activity itself that underlies the major differences in disease phenotype, but rather defective homomeric TASK channels that dominate the unique features of each disorder.

Our results also shed further light on the structural and molecular mechanisms by which one of these pathogenic variants (L122V) may affect TASK channel activity. Their GoF effect is a common feature and such overactive channels clearly have the ability to dampen neuronal excitability. It is also well known that inappropriate spatiotemporal K^+^ channel expression can adversely affect the development of many cell types^37^, especially neurons and both *KCNK3* and *KCNK9* are expressed at early stages in the developing CNS^38^. However, not all of the DDSA associated variants result in a major increase in whole-cell currents and in KIS many of the variants result in reduced TASK-3 channel activity. Therefore, it may be the common loss of sensitivity to GqPCR-mediated inhibition found in all these variants which is just as significant in defining their pathogenic phenotype. Such a regulatory defect may be especially important in the respiratory phenotype seen in all DDSA probands and in some examples of KIS because GqPCR-mediated inhibition of TASK channels contributes to depolarisation of cells in the carotid body as well as in pacemaker neurons in the brainstem important for sleep/wakefulness transition and respiratory rhythmogenesis. Abnormal respiratory control would therefore be expected to result from such defective channel regulation, and the maternal imprinting of *KCNK9* leading to mosaicism could explain why only a subset of KIS probands are reported to have sleep apnea, compared to its defining presence in all known DDSA probands. But there is also precedent in other genetic conditions involving defective K^+^ channels where, paradoxically, both LoF and GoF variants lead to neuronal hyperexcitability^39^ and/or neurodevelopmental disorders^37^. Consequently, much work remains to be done to in order to understand this complexity.

Our cryoEM structure of the L122V variant at the TM2.6 site in TASK-3 also reveals how even subtle changes in structure can have dramatic functional consequences. Although the hydrophobicity of this side chain does not change, the slight increase in volume of the inner cavity just below the filter may be enough to affect a critical step in permeation *i.e*., dehydration of the K^+^ ion prior to its entry into the filter and is also consistent with the activatory effect of hydrophilic substitutions at this site^34, 35^. Indeed, recent detailed simulations of K^+^ permeation in open state conformations of other K2P channels has shown that K^+^ permeation through the filter is tightly coupled to gating^40^, however, open state structures of TASK channels are not currently available. This mutation therefore highlights the complexity of understanding the effect of missense variants on channel activity and the relationship to clinical phenotype, even when detailed functional assays and structural information are available.

In conclusion, we have characterised the functional and structural properties of a range of novel GoF missense disease-causing variants in TASK channels to expand the molecular basis of inherited TASK channelopathies. These variants also provide insight into the relative contributions of homomeric versus heteromeric TASK channels in their respective disorders. Overall, our characterisation of the structural and functional effects of these pathogenic variants not only has important implications for understanding genotype/phenotype correlations, but also future drug development studies for both these rare channelopathies and other more common disease states involving TASK channels.

## Acknowledgments & Funding Information

This work was directly supported by grants from the Biotechnology and Biological Sciences Research Council and Medical Research Council to S.J.T. (BB/T002018/1, BB/S008608/1, and MR/W017741/1). It was also supported by the Wellcome Trust as part of the OXION Initiative in Ion Channels and Membrane Transport in Health and Disease (WT084655MA and 102161/B/13/Z). K.M.C. was funded by the UKRI-BBSRC Interdisciplinary Bioscience Doctoral Training Partnership (BB/M011224/1). We are also grateful for the important support from patients and families, our UK and international collaborators, brainbank and biobanks, and grateful for other essential funding from the MSA Trust, UK Dementia Research Institute, National Institute for Health Research University College London Hospitals Biomedical Research Centre NIHR-BRC), Michael J Fox Foundation (MJFF), Fidelity Foundation, Rosetrees Trust, EAN, ERDERA: European Union’s Horizon Europe research and innovation programme, Dolby Family Fund, Alzheimer’s Research UK (ARUK), Mission MSA, Parkinson’s disease UK, Parkinson’s Foundation, Muscular Dystrophy UK, Ataxia UK, CureDRPLA, ALS Association, National Ataxia Foundation (NAF), Target ALS Foundation, Medical Research Foundation and the National Brain Appeal. JF and CFW were supported by the National Institute for Health and Care Research Exeter Biomedical Research Centre. The views expressed are those of the authors and not necessarily those of the NIHR or the Department of Health and Social Care.

## Author contributions

K.M.C., R.M. and S.J.T. conceived/designed the principal elements of the study. All authors generated, analyzed or interpreted data, or provided clinical data. T.J-T, S.J.T, and K.M.C. drafted the manuscript and all authors contributed to the final version.

## Data Availability

All data produced in the present study are available upon reasonable request to the authors

---

**Supplementary Figure 1:**
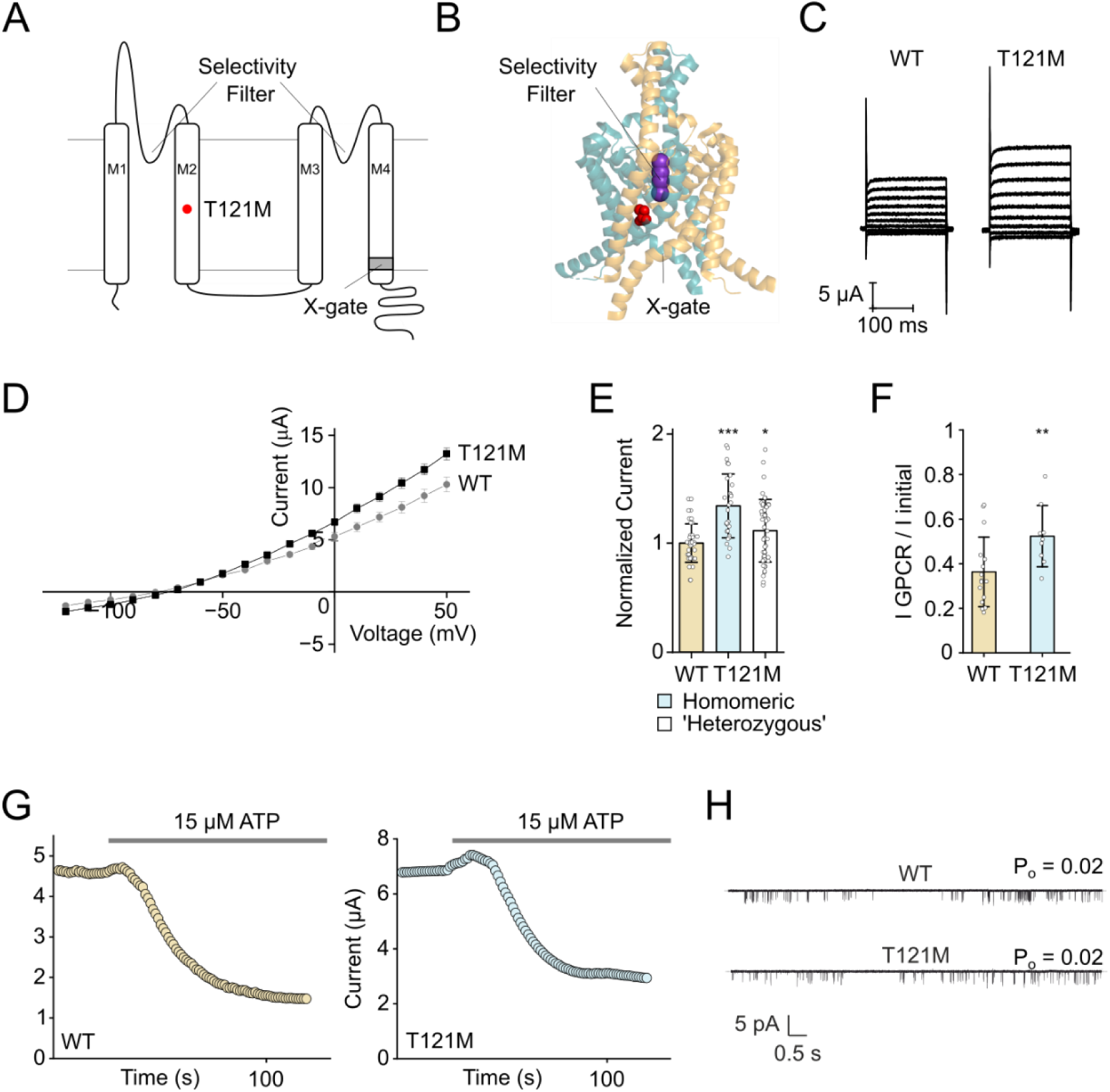
Functional effects of the T121M mutation. (**a**) 2D topological model of a TASK-1 subunit with the position of the T121M mutation labelled in red. The X-gate is labelled in dark grey. (**b**) TASK-1 crystal structure (PDB: 6RV2). Subunits are shown in teal and orange respectively. The T121M mutation is shown as red spheres. (**c**) Representative TEVC recordings of WT TASK-1 and T121M mutant currents in response to voltage steps from −120 to +50 mV in 20 mV steps from a holding potential of −80 mV. (**d**) Current–voltage plot of WT TASK-1 (*n* = 12) and T121M (*n* = 16); data are presented as mean ± s.e.m. (**e**) Currents for homomeric and ‘heterozygous’ T121M channels formed from 1:1 coexpression of WT TASK-1 and T121M normalized to WT current at +50 mV: WT (*n* = 27), T121M (*n* = 28), T121M-WT (*n* = 48) data are presented as mean ± s.d. (**f**) Residual current for homomeric T121M after addition of 15 μM ATP. WT TASK-1 (*n* = 15), T121M (*n* = 9); data are presented as mean ± s.d. * = P<0.05, ** = P<0.01, *** = P<0.001, two-tailed Student’s *t*-test. (**g**) Representative currents at +50 mV of WT TASK-1 and T121M channels, over time while adding 15 µM ATP. (**h**) Single-channel *Po* values at –100mV, WT (*n* = 8) and T121M (*n* = 8).

**Supplementary Figure 1:**
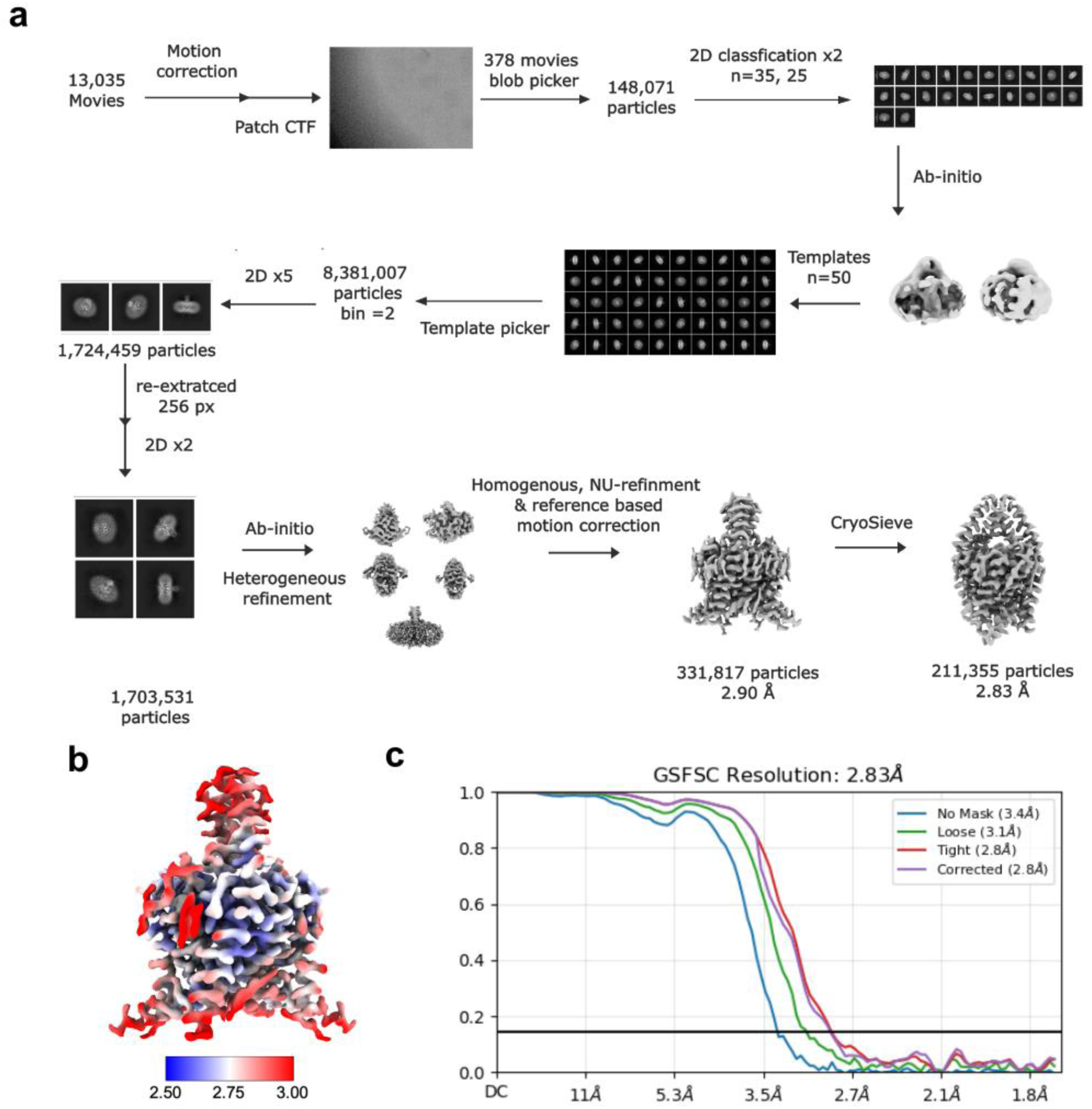
Determination of CryoEM Strutcure of the L122V mutation. (**a**) Image processing workflow for TASK-1 L122V. (**b**) Local-resolution of reconstructed map as determined within cryoSPARC (c), Gold-standard FSC curve used for global-resolution estimates within cryoSPARC.

**Table S1.**
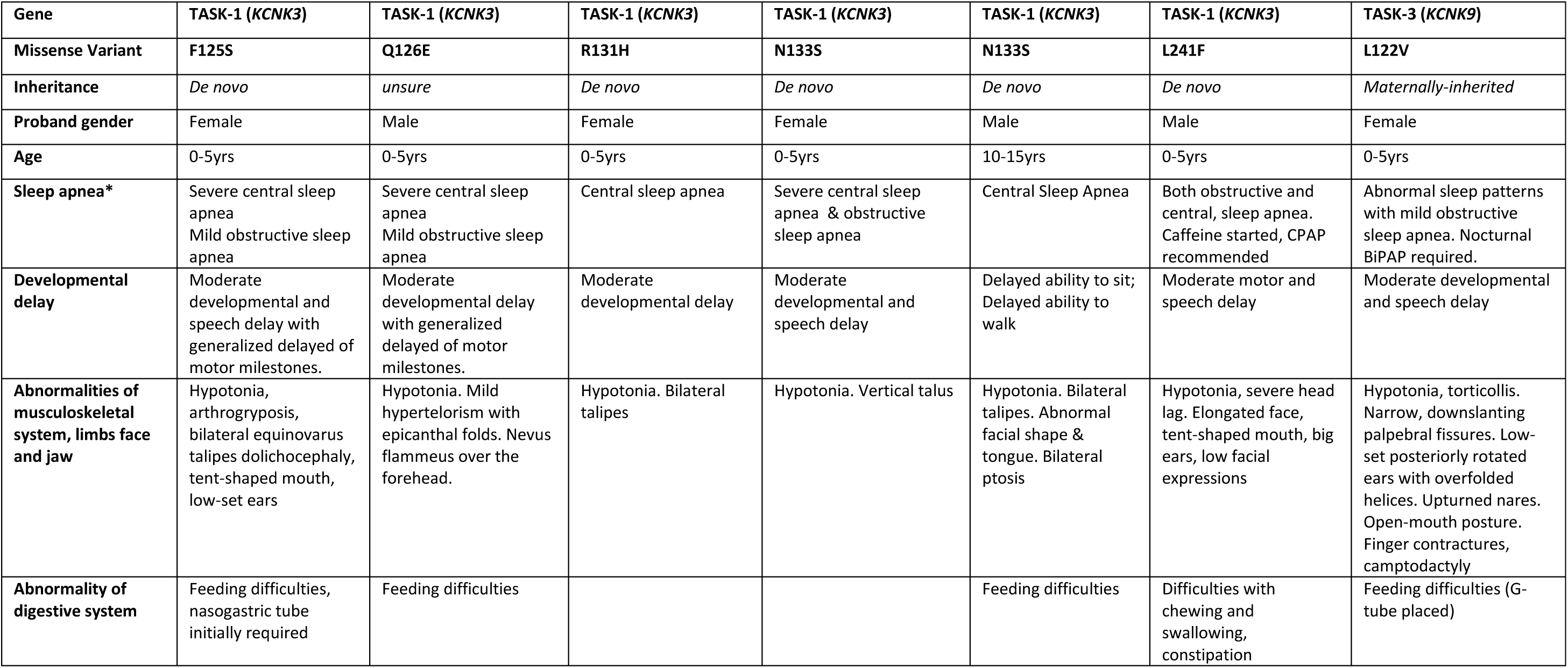

